# Mutation-specific SARS-CoV-2 PCR Screen: Rapid and Accurate Detection of Variants of Concern and the Identification of a Newly Emerging Variant with Spike L452R Mutation

**DOI:** 10.1101/2021.04.22.21255574

**Authors:** Huanyu Wang, Sophonie Jean, Richard Eltringham, John Madison, Pamela Snyder, Huolin Tu, Daniel M. Jones, Amy L. Leber

## Abstract

The emergence of more transmissible and/or more virulent SARS-CoV-2 variants of concern (VOCs) has triggered intensive genomic surveillance, which is costly and difficult to sustain operationally over the long-term. To address this problem, we developed a set of four multiplex mutation-specific PCR-based assays with same-day reporting that can detect five VOCs and three variants of interest (VOIs), as defined in the March 2021 guidelines from the United States (US) Centers for Disease Control and Prevention. The screening results were compared to the whole genome sequencing (WGS) and showed 100% concordance for strain typing for B.1.1.7 (25) and P.1 (5) variants using Spike (S) mutations N501Y, E484K and H69_V70del assays. The S L450R assay, designed to detect the B.1.427/429 VOCs, also identified multiple isolates of a newly emerging multiply-mutated B.1.526.1 variant that is now rapidly increasing in the Eastern US. PCR approaches can be easily adopted in clinical laboratories, provide rapid screening methods to allow early detection of newly emergent variants and to efficiently triage cases for full genomic sequencing.

## Introduction

The novel severe acute respiratory syndrome coronavirus type 2 (SARS-CoV-2) has evolved considerably in the last six months. Most mutations of interest occur in the Spike (S) protein, the viral protein that binds to the Angiotensin Converting Enzyme 2 (ACE2) cell receptor to initiate the attachment of the virus. Beginning in late 2020, several biologically significant S mutations were shown to be associated with increased transmissibility and virulence, diminished protection by antibodies from convalescent or vaccinated anti-sera as well as decreased response to monoclonal antibody treatment. Therefore, strategies to cost-effectively monitor for shifts in SARS-CoV-2 variants are needed.

Three variants of concern (VOCs) with multiple mutations in the S gene have been identified as particularly concerning. The B.1.1.7 (501Y, V1) variant emerged in England and rapidly became the dominant variant in the UK and has now spread to more than 50 countries [1]. B.1.1.7 contains 8-13 different S mutations, including, N501Y in the receptor binding domain (RBD). Studies suggested that it may be associated with higher transmissibility and increased virulence [2, 3]. The B.1.3.5.1 (510Y. V2) variant was first reported in late 2020 in South Africa [4] and variant P.1 (20J/501Y.V3) arose in November 2020 in Brazil. The three spike mutations (K417N/T, E484K and N501Y) carried by both the Brazil and South African variants are associated with increased binding to the human ACE 2 receptor, are more transmissible and mediate partial escape from the protective humoral immunity resulting from prior infection or vaccination [5-7].

In March 2021, the US Centers for Disease Control and Prevention (CDC) included B.1.427/B.1.429 as an additional VOC due to its rapid rise in incidence, and introduced the “variant of interest” (VOI) category to encompass strain mutations that might override neutralizing antibody responses, principally S E474K [8, 9]. B.1.427/B.1.429 carries the S L452R mutation which appears linked to the increased transmissibility of this strain [10]. S L452R may be associated with reduction in neutralization using convalescent, post-vaccination sera and resistance to the neutralization by some monoclonal antibodies that have acquired emergency use authorization for treatment [10-12].

The emergence of these VOC highlights that enhanced surveillance is urgently needed. Real time identification of both VOC and VOI will have significant impact not only on management of this on-going pandemic but also patient care. However, this is not possible without local sequencing capacity, which is not always available or may be limited in throughput capacity; WGS is also costly and takes considerable time to complete. An accurate screening strategy would allow the selective use of WGS by targeting samples of interest. This would maximize the use of WGS and broaden the clinical laboratories able to actively participate in identification of VOC.

Here, we developed reverse-transcriptase real time PCR (rt-PCR) based assays to screen for spike protein deletions 69-70, 242-244 and mutations N501Y, E484K and L452R in clinical samples known to be positive for SARS-CoV-2. The pattern of positivity accurately typed each VOC (as confirmed by genomic sequencing) except for B.1.427/429 where the majority of cases detected (in March-April 2021) corresponded to a newly emerging multiple mutated variant (previously types as B.1.526.1).

## Materials and Methods

### Study Samples

Nasopharyngeal swab samples were collected from pediatric patients and employees of Nationwide Children’s Hospital, Columbus, Ohio. A flocked NP swab was used and placed into viral transport media for transport to the laboratory for testing by a SARS-CoV2 PCR assay. This surveillance study utilized residual sample for assay validation and was thus exempt human subject research. The set comprised 247 samples, including 156 SARS-CoV-2 positive swabs from February 2^nd^ to April 1^st^ 2021 that included all adequate samples with Ct values less than 35, and 91 samples collected from Jan 1^st^ 2021 to Feb 1^st^ 2021 with Ct values less than 35 that were randomly selected from adequate residual samples.

### Diagnostic SARS-CoV-2 PCR Assay

We utilized a laboratory-developed modification of the CDC SARS-CoV-2 rt-PCR assay which received Emergency Use Authorization by the FDA on April 17, 2020. (https://www.fda.gov/media/137424/download; accessed March 24, 2021) Briefly, total nucleic acid was obtained by extraction using the NucliSENS easyMag platform (bioMerieux, Durham, NC) and 5μL of the eluate was added to a 25 μL total volume reaction mixture [5µl of 1XTaqPath 1-Step RT-qPCR Master Mix, CG (Thermo Fisher Scientific, Waltham, MA), 1.5 µl of RT-PCR primer/probe set (N1 or N2, 2019-nCoV kit, Integrated DNA Technologies, Coralville, IA) and 13.5µl nuclease-free water]. The RT-PCR was carried out using the Applied Biosystems™ QuantStudio™ 7 Flex Real Time PCR detection system with QuantStudio™ Real-Time PCR software v.1.3 (Thermo Fisher Scientific, Waltham, MA) with the following running conditions: 25°C for 2min, 50°C for 15 minutes, enzyme activation at 95°C for 2min, and 45 cycles of 95°C for 3 sec and 55°C for 30 sec.

### Mutation Screening Assays

SARS-CoV-2 positive sample were screened by four multiplex rt-PCR assays, with results available same day.

The sequences for detection of Δ69-70 were adapted from a multiplex real-time RT-PCR assay for detection of SARS-CoV-2[13]. The probe overlaps with the sequences that encode amino acids 69-70, therefore, a negative result for this assay predicts the presence of a detection S 69-70 in the sample. Using a similar strategy, a primer/probe set that targets the detection S Δ242-244 was designed and was run in the same reaction with S Δ69-70. In addition, three separate assays were designed to detect spike mutations 501Y, 484K and 452R and wild types 501N, 484E and 452L. The sequences for each oligonucleotide are presented in Table 1.

**Table 1:**
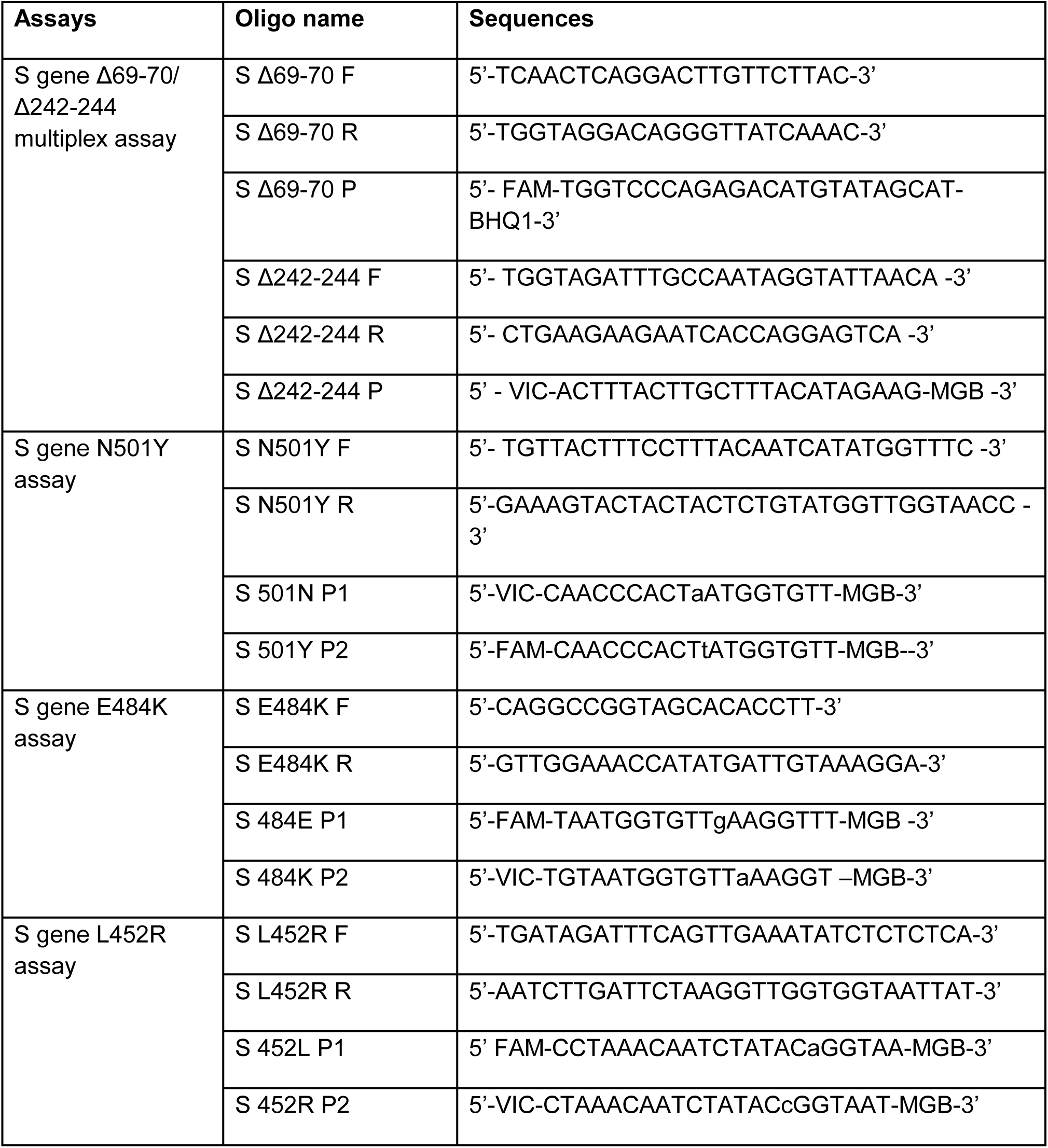
Mutation-specific reverse-transcriptase PCR: primers and probes.

Briefly, 5μL of the total nucleic acid eluate was added to a 20 μL total volume reaction mixture [1XTaqPath 1-Step RT-qPCR Master Mix, CG (Thermo Fisher Scientific, Waltham, MA), 0.9μM of each primer and 0.2μM of each probe]. The RT-PCR was carried out using the ABI 7500 thermocycler (Life Technologies, Grand Island, NY). The N501Y, E484K and L452R assays were carried out with the following running conditions: 25°C for 2 min, then 50°C for 15 min, followed by 10 min at 95°C, 45 cycles of 95°C for 15s and 65°C for 1 min. The Δ69-70/Δ242-244 assay was run under the following conditions: 25°C for 2 min, then 50°C for 15 min, followed by 10 min at 95°C, 45 cycles of 95°C for 15s and 60°C for 1 min.

Samples that yielded negative result(s) in Δ69-70/Δ242-244 assay or were positive for 501YP2, 484KP2 and L452RP2 were considered screen-positive and assigned to a VOC based on the scoring in Table 2. All screening positive samples and a similar number of screening negative samples collected in the same period were sent for WGS.

**Table 2:** Strain-typing rules for interpretation of screening results.

|  | Mutations |  |  |  |  |
| --- | --- | --- | --- | --- | --- |
|  | Δ69-70 | Δ242-244 | N501Y | E484K | L452R |
| **Variants of Concern** |  |  |  |  |  |
| B.1.1.7 | Pos | Neg | Pos | Neg | Neg |
| B.1.351 | Neg | Pos | Pos | Pos | Neg |
| P.1 | Neg | Neg | Pos | Pos | Neg |
| B.1.427/ B. 1.429 or B.1 80G-157L-4 | Neg | Neg | Neg | Neg | Pos |
| **Variants of Interest** |  |  |  |  |  |
| B.1.525 | Pos | Neg | Neg | Pos | Neg |
| R.1, P.2, B.1.525, B.1.526, | Neg | Neg | Neg | Pos | Neg |
| B.1.2/501Y or B.1.1.165 | Neg | Neg | Pos | Neg | Neg |

### SARS-CoV-2 Whole Genome Sequencing

Previously extracted RNA underwent 1st and 2nd strand cDNA synthesis (NEBNext® Ultra# II Non-Directional RNA Second Strand Synthesis Module, NEB, Ipswich, MA) followed by sequencing using two different clinically validated amplicon-based methods. Most samples were analyzed using the SARS-CoV-2 Research Panel primers (ThermoFisher) on the Ion Chef-S5 sequencer (ThermoFisher) per manufacturer’s conditions. The remaining sequences were analyzed using the CovidSeq kit (Illumina, San Diego, CA) per manufacturer’s conditions and sequenced on the NextSeq 550 sequencer (Illumina). Strain typing on the two different assays were cross-validated using a set of 20 SARS-CoV-2 positive and 5 SARS-CoV-2 negative samples.

### Genomic Analyses and Strain Typing

Sequence analysis tools include custom pipelines utilizing GATK and Mutect2 (Broad Institute) and Dragen SARS-COVID variant detection (for CovidSeq assay). Adequate sequencing required coverage of strain-distinguishing areas of the ORF1a, S, N and ORF8 genes at least 100X depth. All but two sequences in this series yielded adequate strain-typable sequences and no sample contained a mixed population of viruses. Viral sequences were strain-typed using Pangolin (https://cov-lineages.org) and NextStrain criteria (https://nextstrain.org/blog/2021-01-06-updated-SARS-CoV-2-clade-naming). Novel combinations of mutations were investigated for similar viruses using the laboratory internal sequence database (1110 sequences as of 4/1/21) and/or the GISAID database (accessed at gisaid.org).

## Results

### Correlation of RT-PCR Mutation Screen Results with Whole Genome SARS-CoV-2 Sequencing

From Jan 2021 to April 1^st^, we screened 247 samples for VOC and VOI. Among all screened samples, 128 samples were collected from hospital employees, 91 samples were from outpatients and 28 samples were from hospitalized or emergency room patients. A total of 102 samples were sent for WGS.

There were 61 screen-positive samples and 41 screen-negative samples sent for WGS. The screening assays were 100% concordant with the WGS for identification of the three RBD mutation (Table 3).The screening assays successfully predicted the presence of all 25 B.1.1.7 and 5 P.1 VOCs (Table 4). Six samples were positive for only the E484K mutation, with five confirmed to be P.2 and one confirmed to be R.1 by WGS. The screening PCR assays had 100% negative agreement when compared to WGS for prediction of these VOCs and VOIs. Sixteen of the samples were positive only for N501Y and were shown by sequencing to be the 20G/501Y strain we have previously described [14] in fourteen and B.1.1.165 in two. Eight samples were screen-positive for S L452R with 3 of them strain typed as B.1.427/B.1.429, and five representing the newly emerging B.1.526.1 variant described in more detail below.

**Table 3:** Performance of the Screening Assays for Detection of RBD Mutations.

|  |  | WGS n=102 |  |  |  |  |  |
| --- | --- | --- | --- | --- | --- | --- | --- |
|  |  | L452R |  | E484K |  | N501Y |  |
|  |  | Pos | Neg | Pos | Neg | Pos | Neg |
| Screening assays | Pos | 8 | 0 | 12 | 0 | 46 | 0 |
|  | Neg | 0 | 94 | 0 | 90 | 0 | 56 |
| PPA % |  | 100 |  | 100 |  | 100 |  |
| NPA % |  | 100 |  | 100 |  | 100 |  |

PPA: positive percentage agreement; NPA: negative percentage agreement

**Table 4:** Performance of the screening assays for detection of variants.

|  |  | WGS n=102 |  |  |  |  |  |  |  |  |  |  |  |  |  |
| --- | --- | --- | --- | --- | --- | --- | --- | --- | --- | --- | --- | --- | --- | --- | --- |
|  |  | Variants of Concern |  |  |  |  |  |  |  | Variants of Interest |  |  |  |  |  |
|  |  | B.1.1.7 |  | B.1.351 |  | P.1 |  | B.1.427 or B.1.429 |  | B.1.525 |  | P.2, B.1.526 or R.1 |  | 20G/501Y as in Ref 14 or B.1.1.165 |  |
|  |  | Pos | Neg | Pos | Neg | Pos | Neg | Pos | Neg | Pos | Neg | Pos | Neg | Pos | Neg |
| **Screening assays** | Pos | 25 | 0 | 0 | 0 | 5 | 0 | 3 | 5<sup>\$</sup> | 1 | 0 | 6* | 0 | 16 | 0 |
|  | Neg | 0 | 77 | 0 | 102 | 0 | 97 | 0 | 94 | 0 | 101 | 0 | 96 | 0 | 86 |
| **PPA %** |  | 100 |  | N/A |  | 100 |  | 37.5 |  | 100 |  | 100 |  | 100 |  |
| **NPA %** |  | 100 |  | 100 |  | 100 |  | 100 |  | 100 |  | 100 |  | 100 |  |

\$All five samples were identified carrying a newly emerged variant: B.1.526.1
\*Five samples were identified as strain P.2 and one sample was strain R.1
PPA: positive percentage agreement; NPA: negative percentage agreement

All negative screening samples were negative for VOC or VOI by WGS. Forty one screen-negative samples were found to be commonly encountered B.1 or B.1.2 with no S mutations of concern. No sample was positive only for S deletion Δ69-70 or S deletion Δ242-244. One sample that had suboptimal amplification in the Δ69-70 assay was found to have a mutation in the primer binding site.

### Timing of Emergence of PCR-detected Variants of Concern Matches Results from

From January to April 1^st^ 2021, we screened 247 SARS-CoV-2 samples. As shown in Figure 1, the earliest case with B.1.1.7 was identified in mid-February 2021, subsequently, cases of B.1.1.7 were found in every batch of samples tested. The earliest case with P.1 was also collected in mid-February 2021, with a substantial increase in P.1 positive cases over time. The first B.1.429 in our population appeared in late March 2021.

**Figure 1.**
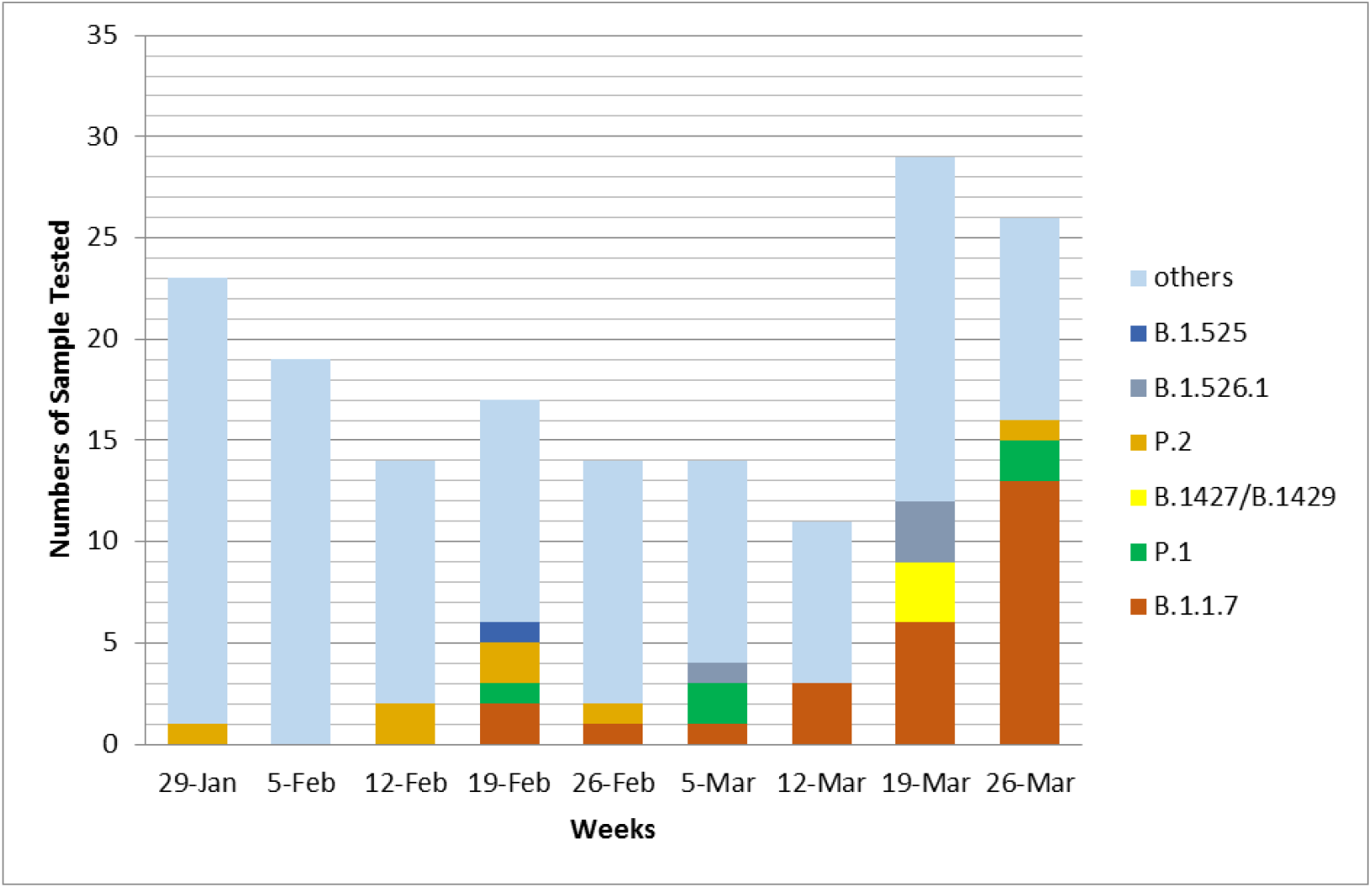
Shifts in the proportions of SARS-CoV-2 strains from January to March 2021.

In the third week of February, 2021, one sample collected from a patient was found to carry both N501Y and E484K mutations and was sent for WGS. The sequences of this sample closely relate to Brazil P.1. VOC and was designated into lineage B.1.1.28 and was the first P.1. sample reported in the state of Ohio (EPI_ISL_1164183). In the same week, another sample collected from an employee was found to carry Δ69-70 and E484K mutation that represent the first Ohio sample of the emerging B.1.525 lineage (EPI_ISL_1203843).

Three of the L452R-bearing viruses in our screen were documented to be B.1.427/429. This VOC first appeared in California in July 2020 (EPI_ISL_765997) and is characterized by Spike mutations S13I, W152C as well as L452R. In early November 2020, B.1.427/429 became the predominant strain in Southern California before spreading throughout the United States in December-January 2020. The observed increase in B.1.427/429 in our study matches that seen in CDC totals from WGS in Ohio.

### Emergence of the B.1.526.1 L452R-containing variant

Unlike the other VOC in our study, we noted that a positive screen for L452R still required sequencing to definitively strain type. This was due to the emergence in late March in our area of another L452R-bearing strain besides the B.1.427/429 VOC. Five of 8 (62.5%) screen-positive L452R-containing viruses from March 2021 represented a newly emerging strain that has been tentatively strain-typed as B.1.526.1 (Table 5). The diagnostic PCR Ct values are presented in Table 6. In addition to L452R, this strain has four additional S mutations, D80G, F157S, T859N and D950H, which include both N-terminal and stalk variants. It shares with B.1.427/429, the 20C/B.1 backbone as indicated by mutations ORF1 T265I, ORF1a A1679S, ORF1a L3201P, ORF1b P314L, ORF3a Q57H and S D614G.

**Table 5:** Spike mutations associated with a newly emerged S L452R-containing variant.

| Amino acid change | cDNA location | # GISAID* | # B.1.526.1 viruses (%) | Domain | Other lineages commonly showing the mutation |
| --- | --- | --- | --- | --- | --- |
| p.Asp80Gly | c.239A>G | 587 | 548 (93.4) | NTD |  |
| p.Phe157Ser | c.470T>C | 661 | 588 (89.0) | NTD |  |
| p.Leu452Arg | c.1355T>G | 12,997 | 608 (4.7) | RBD | B.1.427/429, A.2.5 |
| c.2576C>A | p.Thr859Asn | 840 | 461 (54.9) | CR | B.1.36.9 |
| p.Asp950His | c.2848G>C | 459 | 419 (91.3) | HR1 |  |
| 4 or 5 of these S mutations |  | 449 | 449 (100) |  |  |

Abbreviations: NTD, N-terminal domain; RBD, receptor binding domain; HR1, heptad repeat 1;
CR: connecting region.
\*Data as of 4/10/21

**Table 6:**
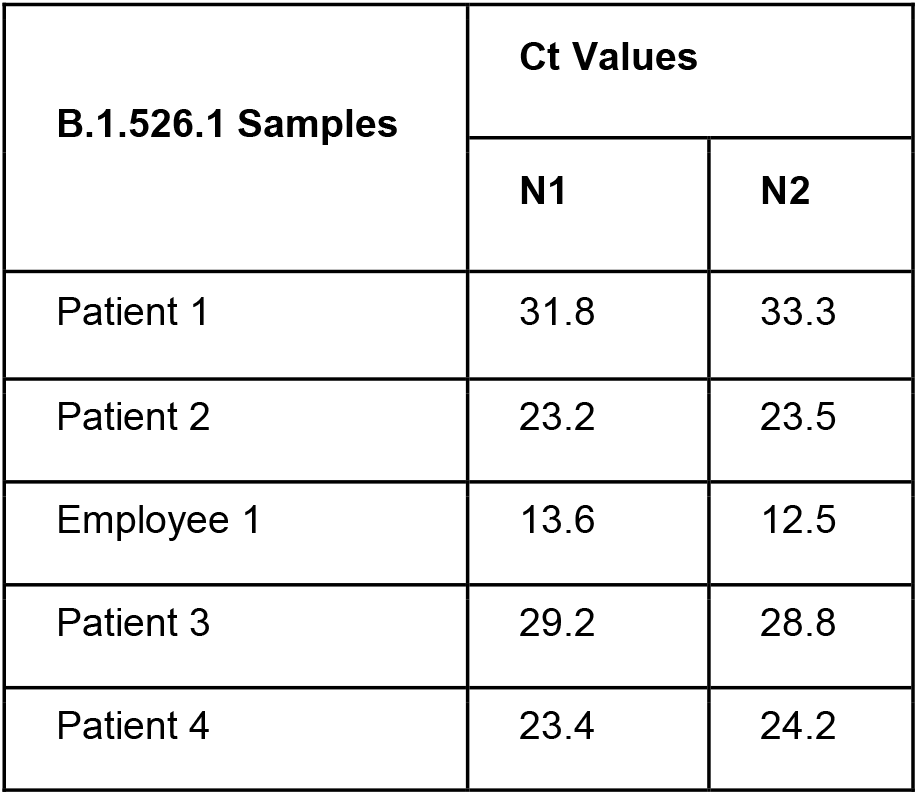
Ct values of the samples carry newly emerged S L452R-containing variant.

| **B.1.526.1 Samples** | **Ct Values** |  |
| --- | --- | --- |
|  | **N1** | **N2** |
| Patient 1 | 31.8 | 33.3 |
| Patient 2 | 23.2 | 23.5 |
| Employee 1 | 13.6 | 12.5 |
| Patient 3 | 29.2 | 28.8 |
| Patient 4 | 23.4 | 24.2 |

Based on the frequency of the Spike mutations in B.1.526.1 viruses reported in GISAID as of April 10 2021, the variant likely developed from a L452R-bearing B.1 strain, then acquired F157S, D80G and Y145del, followed by T859 and D950H. The few GISAID sequences that do not follow this pattern are reported as suboptimal quality studies. A small number (n=7) of B.1.526.1 viruses contain D80N/N81Y instead of D80G. The earliest GISAID-reported instances of B.1.526.1 were two sequences from New York City collected on 12/18/20 (EPI_ISL_794288 and EPI_ISL_794289), with all sequences through 2/5/21 reported from New York City (mostly, Bronx county). Spread to 11 other states (including Ohio) occurred in February and then throughout the United States in March, 2021. Given that 578 of 2369 (24.4%) viruses typed as B.1.526.1 lineage in GISAID do not contain any of the above Spike mutations, lineage classification may be provisional.

A comparison of the incidence of the B.1.427/429 and B.1.526.1 strains in GISAID during 2021 shows a declining proportion of the forms and increases in the latter in March matching our observations in Columbus (Figure 2).

**Figure 2.**
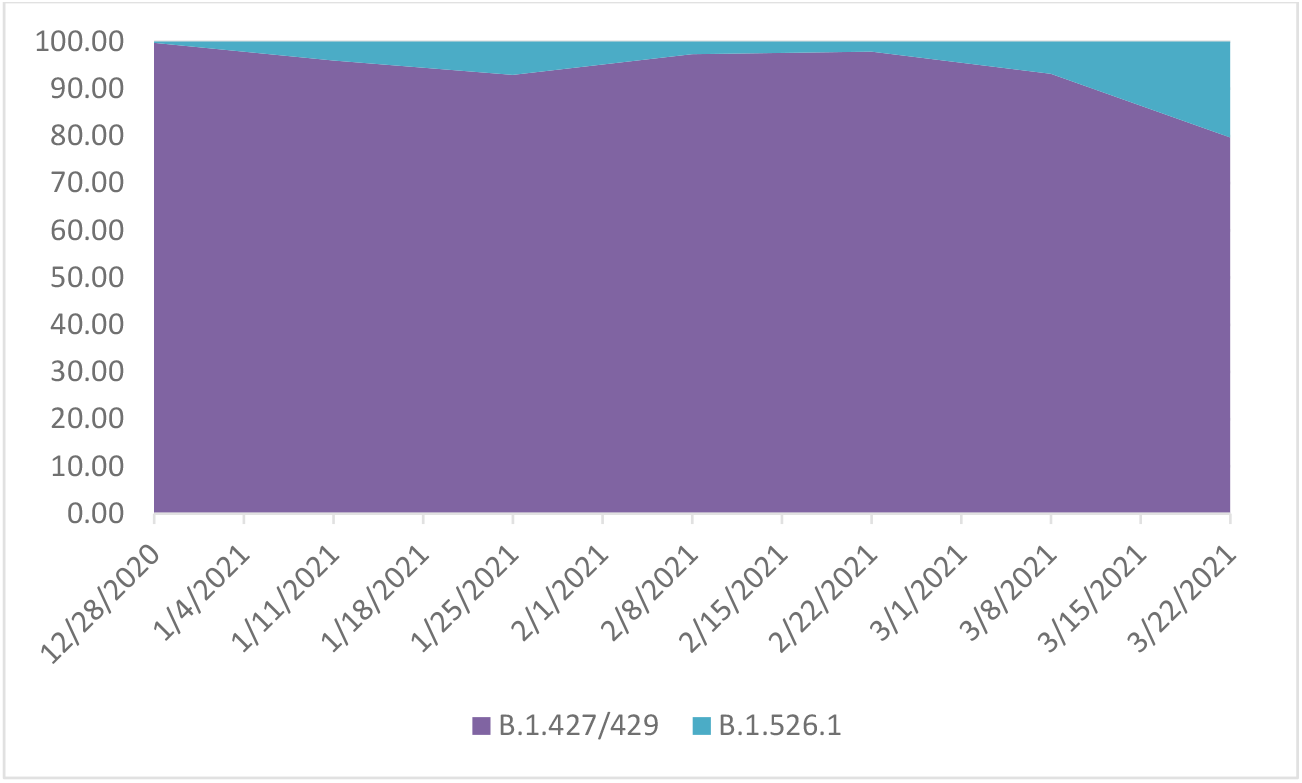
Comparison of incidence in the United States of B.1.427/429 and B.1.526.1 in 2021. Spike L452R-bearing SARS-CoV-2 genomes from North America were downloaded from GISAID and strain type confirmed by mutation pattern. Graphed is the proportion of B.1.427/429 versus B.1.526 viruses during each 2-week period in 2021.

## Discussion

We describe a multiplex, mutation-specific rt-PCR-based strategy to rapidly and reliably screen for SARS-CoV2 genetic variants in a clinical laboratory setting. These assays detect all three target RBD spike mutations and have a 100% concordance with the strains identified by WGS. Using simple rules based on the combination of assay results, the strategy successfully identified all B.1.1.7 and P.1 samples, with the frequencies of each of the VOCs detected matching their emergence by the CDC and Department of Health WGS surveillance studies over the last several months. Using this strategy, we successfully identified and reported the first P.1 variant in Ohio from a patient which suggested that the P.1 variant has already spread in the community; a finding confirmed in subsequent reports.

However, for S L452R-positive screens, we noted that both B.1.427/429 VOC and a newly emerging B.1.526.1 strain were detected [15]. The latter samples were identified only in the latest March 2021 samples and match the observed spread of GISAID-reported B.1.526.1 viruses outside of the state of New York. Although both of these strains show multiple mutations in other areas of the Spike gene, the L452R mutation is their only RBD mutation. This mutation has been associated with resistance to neutralization by RBD-binding monoclonal antibodies, such as bamlanivimab [12], which has affected the use of these therapies. The single RBD mutation in these two strains is in contrast to other VOC which besides N501Y and E484K (in two of them, P.1 and B.1.351) show additional mutations known to alter receptor binding affinity [16-18]. It will be important to monitor sequential samples from these strains to see if the acquire additional RBD mutations, particularly at antibody escape residues, as B.1.1.7 viruses have done in some cases.

Mutation-specific PCR assays can be run in real-time with a turn-around time of several hours, which is significantly faster than WGS and in this configuration can be performed cost-effectively on all PCR-positive samples. This broader approach is clearly superior to the use of other VOC indicators such as S-gene dropout in the 3-target diagnostic RT-PCR assay (TaqPath kit, Thermo Fisher Scientific, Waltham, MA) which has been reported by others as a method to detect the H69-V70del associated with B.1.1.7 [9, 19-22]. Similarly, PCR-based N501Y and E484K mutation-specific assays are sensitive for detection of P.1 and B.1.351 [23, 24], respectively, but will not definitively distinguish them as an increasing number of VOIs also bear these mutations. In this study, we could not fully evaluate all scoring combinations, particularly the use of co-positivity for N501Y and Δ242-244 as a marker for the B.1.351 VOC, as we did not find any examples of this virus in our test population. However, the combination of assay presented here has more diagnostic utility in distinguishing VOC and VOI than single mutation-specific PCR approaches.

There are some limitations to a reflex mutant-PCR panel strategy. First, many diagnostic PCR platforms can deplete swab material, leaving an inadequate volume of residual sample for a multi-tube mutation screen. These assays also did not perform well when the diagnostic PCR Ct value is greater than 35 cycles. However, this problem is shared with amplicon-based WGS [25]. As demonstrated by our finding with L452R-positives, new variants of SARS-CoV-2 are still emerging so confirmation of some results by WGS will still be required. Finally, to evaluate for novel mutations, a sampling of screen-negative specimens for WGS will be an important component of a full surveillance strategy.

In summary, the use of a panel of multiplex, mutant-specific rt-PCR assays represents an ideal balance of cost, turn-around-time and accuracy for detecting SARS-CoV-2 VOC and VOI. The technology is amenable for use in a larger number of clinical laboratories than next-generation sequencing. In our current environment requiring rapid strain typing to guide both treatment decisions and public health measures, such a rapid and accessible approach will be essential.

## Data Availability

All data is available for review.

## Acknowledgments

For the GIS sequences in Figure 2, we acknowledge the originating laboratories responsible for obtaining the specimens and the submitting laboratories where genetic sequence data were generated and shared via the GISAID Initiative.

